# Risk factors associated with Ahmed valve implant dysfunction in patients with neovascular glaucoma during the first postoperative year

**DOI:** 10.1101/2022.05.10.22274303

**Authors:** K. Ureña-Wong, J. Jimenez-Román, R. Rivas-Ruiz, J A. Morales-Gonzalez

**Affiliations:** Asociacion para evitar la ceguera en Mexico; Instituto Mexicano del Seguro Social; Instituto Politecnico Nacional

**Keywords:** Neovascular glaucoma, Ahmed valve, risk factors, survival analysis

## Abstract

**Background:** The current surgical management of Neovascular Glaucoma (NVG) is with the placement of the Ahmed Valve. However, up to 70% fail and the factors associated with this failure are unknown. Recognizing the factors associated with dysfunction will help to identify patients prone to failure promptly to establish prompt management and improve visual and functional prognosis.

**Objective:** To determine the risk factors associated with Ahmed valve dysfunction in patients with NVG during the first year after surgery.

**Population:** Patients diagnosed with VNG who have undergone Ahmed valve implantation in the Glaucoma - APEC service, during the 2013 to 2019 period.

**Methods:** ambispective cohort. Patients with NVG defined as the Presence of neovessels in the iris or angle of the anterior chamber were included, those who had a history of implantation of a drainage device, and patients with a diagnosis of glaucoma in the terminal phase, worse visual acuity of perception were excluded. of light. The research unit was eyes subjected to Ahmed valve implantation. The demographic characteristics of the patients were evaluated, as well as the characteristics of the eyes.

The primary outcome and the secondary outcomes were adverse events, visual acuity, and final intraocular pressure. An unadjusted Cox proportional hazards model was used to determine risk factors such as Hazzard ratios (HR). Those significant factors were included in a Cox proportional hazards model to adjust for the main confounding variables.

**Results:** 174 eyes were included. Baseline intraocular pressure (IOP) was 46.03 (±11.8) mmHG. Baseline VA in LogMAR was 1.52(0.89-2.3). Risk factors were: Age <50 years with HR 1.54(95% CI 1.04-2.30); HB1Ac >8% HR 1.71 (95% CI 1.12-2.60) and presence hypertensive phase HR 3.13 (95% CI 1.57-6.23). The multivariate model was adjusted for the following variables, baseline IOP less than 40mmHg HR 1.60 (95% CI 1.04-2.47); HB1Ac >8% HR 1.80 (95% CI 1.16-2.78); FP7 valve type HR 1.75(95% CI 1.04-2.94) and presence of hypertensive phase HR 3.24 (95% CI 1.60-6.59).

**Conclusion:** Basal IOP less than 40mmHg, HB1Ac >8%, type of FP7 valve, surgery performed by a resident, lack of photocoagulation after implantation, and the presence of a hypertensive phase are the independent risk factors for implant valvular dysfunction.

## Background

Neovascular glaucoma (NVG) is secondary glaucoma that is difficult to control. It is characterized by elevated IOP about the presence of neovessels and extracellular matrix tissue in the iris, angle, and anterior chamber.(1). This type of glaucoma represents 3.9% of all glaucomas. (2). NVG can occur in 2.1% of all diabetic patients. In diabetic patients with proliferative retinopathy, NVG occurs in 21.3% of cases (3). Diabetic patients have a 33% risk of developing NVG in the fellow eye(4)(5).

Surgical management of neovascular glaucoma consists mainly of filtering surgeries, that is, implantation of drainage devices. However, in patients with NVG, the failure rate of these devices is higher than in patients without NVG (success rate at 1 year = 89.2% versus 73.1% (p = 0.0096)(6)

The use of drainage devices is preferred in situations where a significant decrease in IOP is required. Some series report a decrease of up to 50% compared to baseline IOP. Souza et al. report that the basal IOP 30.4±10.7 mmHg had a post-surgical decrease to 17.0±5.0 mmHg at 12 months and 15.9±3.0mmHg at 60 months of follow-up (p<0.05).(7). When comparing Ahmed valve implant (AVI) with trabeculectomy, several studies report that there is no significant difference in final IOP at one year of follow-up, however, IVA has a higher success rate (83.6% versus 88.1%), as well as less need of hypotensive drugs at three and six months after the procedure(8,9).

The survival of these implants has been described from 49% to 70% at one year of follow-up (10). Regarding factors associated with the failure of AVI, the reported results are not very consistent. Hernandez-Oteyza et al. did not find statistically significant differences in the rate of success in patients with IOP <21 mmHg (OR = 1.45, CI = 0.37-6.34, p = 0.76). However, they report that the presence of a hypertensive phase (IOP >21mmHg) at any time during the first year of follow-up is a risk factor for failure of the drainage device (OR = 5.15, CI = 1.49-20.15, p = 0.004)(11). In contrast, Lee, C. et al. performed a Cox proportional hazards model that indicated that eyes with high preoperative IOP had a higher risk of failure RR 1.2 CI95% 1.02-3.54 (p = 0.029)(12).

Sahyoun et al, in a 5-year follow-up, report that a history of Diabetes Mellitus, previous trabeculectomy, or cataract surgery was not associated with an increased risk of failure (p = 0.35, p = 0.31, and p = 0.61, respectively).(13). These results are contrary to what was found by Souza et al. In the Cox proportional hazard model, eyes that had undergone previous glaucoma surgery had a higher risk of failure OR: 3.07 95% CI 1.42-6.66 (p = 0.004)(14).

Due to all of the above, the factors associated with failure could not be established, either due to inadequate assemblies or statistical analyzes without sufficient power.

## Methods

This is an ambispective cohort. Patients diagnosed with NVG who have undergone Ahmed valve implantation in the Glaucoma-APEC service, during the 2013 to 2019 period. Patients with NVG defined as the Presence of neovessels in the iris or angle of the anterior chamber were included, those who had a history of implantation of a drainage device, and patients with a diagnosis of glaucoma in the terminal phase, worse visual acuity of light perception were excluded. The research unit was eyes subjected to Ahmed valve implantation. The demographic characteristics of the patients were evaluated, as well as the characteristics of the eyes.

The sample size was calculated with the difference of proportions formula (15). In the literature, there are several reports of the percentage of survival (time to dysfunction) of this type of implant. Patients who have NVG have a failure rate of 78%, in contrast, those patients who do not have NVG have a failure rate of only 53%, a total of 128 patients is obtained as the total sample size, requiring 64 patients by group to ensure adequate statistical power.

The primary outcome and the secondary outcomes were adverse events, visual acuity, and final intraocular pressure.

For the quantitative variables, the type of distribution was determined through mental methods (Asymmetry and Kurtosis), as well as utilizing the Kolmogorov-Smirnov test. The variables that resulted from free distribution, were represented as median and interquartile range (IQR). The variables that had a normal distribution were summarized as mean and standard deviation.

The bivariate analysis was carried out with Pearson’s X2 test for those dichotomous variables. If expected values were found to be less than five, Fisher’s exact test was used. For dichotomous variables with 3 groups, the linear trend X2 test was used. In the case of quantitative variables, means were compared with the Student’s t - test for independent samples or the Mann-Whitney U test depending on their type of distribution.

The risk factors were calculated from an unadjusted Cox proportional hazards model. Subsequently, for the multivariate analysis, the variables that were significant in the bivariate model were analyzed using Cox proportional hazards to obtain the independent variables associated with the dysfunction of drainage devices.

To determine the median survival time of the drainage devices, a Kaplan-Meier analysis was performed with the construction of survival curves. Both groups were compared using the Log Rank test in the survival curves.

Ethics committee/IRB of Asociación para evitar la ceguera en México gave ethical approval for this work under the protocol number GL-20-02.

The statistical program SPSS (IBM Corp. Released 2020. IBM SPSS Statistics for Windows, Version 27.0. Armonk, NY: IBM Corp) was used and GraphPad Prism version 8.0.0, was used to build the graphs, GraphPad Software, San Diego, California USA.

## Results

A total of 174 eyes of 168 patients were included. The baseline characteristics of the population are presented in Table I. The mean age of the patients was 54.5 years old with an SD of ±9.4 years. 43.1% (75) of the patients were female. A total of 96.6% (168) of the patients had a history of diabetes mellitus, with a median of 16 years of diagnosis and an interquartile range (IQR) of 13 to 20 years. These patients presented a median glycosylated hemoglobin (HbA1c) of 8.37% with an IQR of 7.18-9.70. Grade 3 NVG occurred in 72.4% (126) of the patients. Baseline VA had a median LogMAR of 1.52 (0.89-2.3). Basal intraocular pressure had a mean of 46.03 mmHg with an SD ±11.8mmHg. The post-surgical hypertensive phase occurred in 79.3% (138) of the patients. A total of 57.5% (100) of the patients presented dysfunction of the Ahmed valve implant.

**Table I.**
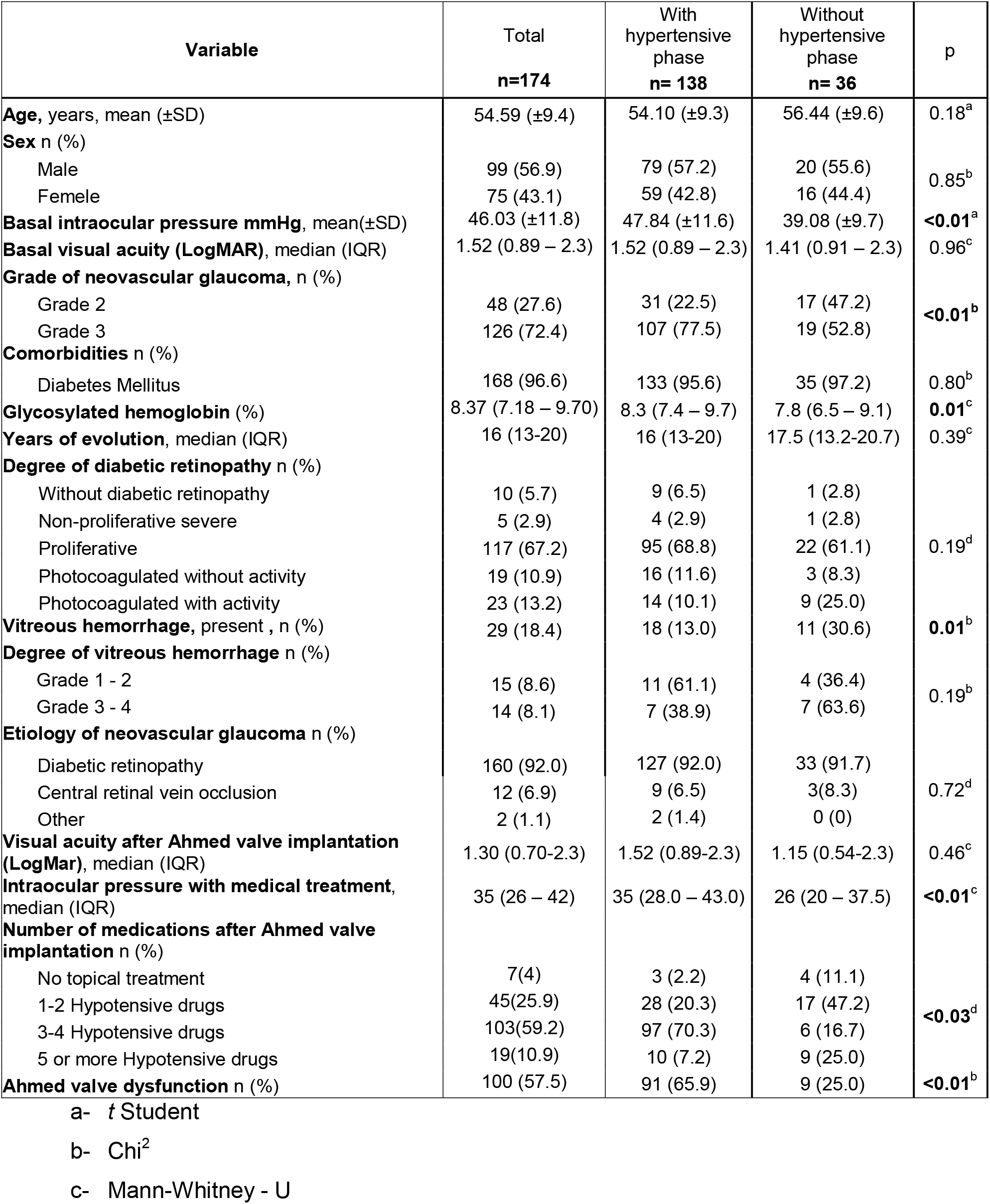
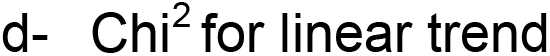
Baseline Characteristics of Patients With Ahmed Valve Implantation.

| Variable | Total<br>n=174 | With<br>hypertensive<br>phase<br>n= 138 | Without<br>hypertensive<br>phase<br>n= 36 | p |
| --- | --- | --- | --- | --- |
| **Age**, years, mean (±SD) | 54.59 (±9.4) | 54.10 (±9.3) | 56.44 (±9.6) | 0.18<sup>a</sup> |
| **Sex** n (%) |  |  |  |  |
| Male | 99 (56.9) | 79 (57.2) | 20 (55.6) | 0.85<sup>b</sup> |
| Female | 75 (43.1) | 59 (42.8) | 16 (44.4) |  |
| **Basal intraocular pressure mmHg**, mean(±SD) | 46.03 (±11.8) | 47.84 (±11.6) | 39.08 (±9.7) | **<0.01<sup>a</sup>** |
| **Basal visual acuity (LogMAR)**, median (IQR) | 1.52 (0.89 – 2.3) | 1.52 (0.89 – 2.3) | 1.41 (0.91 – 2.3) | 0.96<sup>c</sup> |
| **Grade of neovascular glaucoma**, n (%) |  |  |  |  |
| Grade 2 | 48 (27.6) | 31 (22.5) | 17 (47.2) | **<0.01<sup>b</sup>** |
| Grade 3 | 126 (72.4) | 107 (77.5) | 19 (52.8) |  |
| **Comorbidities** n (%) |  |  |  |  |
| Diabetes Mellitus | 168 (96.6) | 133 (95.6) | 35 (97.2) | 0.80<sup>b</sup> |
| **Glycosylated hemoglobin (%)** | 8.37 (7.18 – 9.70) | 8.3 (7.4 – 9.7) | 7.8 (6.5 – 9.1) | **0.01<sup>c</sup>** |
| **Years of evolution**, median (IQR) | 16 (13-20) | 16 (13-20) | 17.5 (13.2-20.7) | 0.39<sup>c</sup> |
| **Degree of diabetic retinopathy** n (%) |  |  |  |  |
| Without diabetic retinopathy | 10 (5.7) | 9 (6.5) | 1 (2.8) | 0.19<sup>d</sup> |
| Non-proliferative severe | 5 (2.9) | 4 (2.9) | 1 (2.8) |  |
| Proliferative | 117 (67.2) | 95 (68.8) | 22 (61.1) |  |
| Photocoagulated without activity | 19 (10.9) | 16 (11.6) | 3 (8.3) |  |
| Photocoagulated with activity | 23 (13.2) | 14 (10.1) | 9 (25.0) |  |
| **Vitreous hemorrhage**, present , n (%) | 29 (18.4) | 18 (13.0) | 11 (30.6) | **0.01<sup>b</sup>** |
| **Degree of vitreous hemorrhage** n (%) |  |  |  |  |
| Grade 1 - 2 | 15 (8.6) | 11 (61.1) | 4 (36.4) | 0.19<sup>b</sup> |
| Grade 3 - 4 | 14 (8.1) | 7 (38.9) | 7 (63.6) |  |
| **Etiology of neovascular glaucoma** n (%) |  |  |  |  |
| Diabetic retinopathy | 160 (92.0) | 127 (92.0) | 33 (91.7) | 0.72<sup>d</sup> |
| Central retinal vein occlusion | 12 (6.9) | 9 (6.5) | 3(8.3) |  |
| Other | 2 (1.1) | 2 (1.4) | 0 (0) |  |
| **Visual acuity after Ahmed valve implantation (LogMar)**, median (IQR) | 1.30 (0.70-2.3) | 1.52 (0.89-2.3) | 1.15 (0.54-2.3) | 0.46<sup>c</sup> |
| **Intraocular pressure with medical treatment**, median (IQR) | 35 (26 – 42) | 35 (28.0 – 43.0) | 26 (20 – 37.5) | **<0.01<sup>c</sup>** |
| **Number of medications after Ahmed valve implantation** n (%) |  |  |  |  |
| No topical treatment | 7(4) | 3 (2.2) | 4 (11.1) | **<0.03<sup>d</sup>** |
| 1-2 Hypotensive drugs | 45(25.9) | 28 (20.3) | 17 (47.2) |  |
| 3-4 Hypotensive drugs | 103(59.2) | 97 (70.3) | 6 (16.7) |  |
| 5 or more Hypotensive drugs | 19(10.9) | 10 (7.2) | 9 (25.0) |  |
| **Ahmed valve dysfunction** n (%) | 100 (57.5) | 91 (65.9) | 9 (25.0) | **<0.01<sup>b</sup>** |

a- t Student
b- Chi<sup>2</sup>
c- Mann-Whitney - U
d- $\chi^2$ for linear trend

The most common etiology of neovascular glaucoma was diabetic retinopathy in 92% (160) of the cases, the other etiologies found were central retinal vein occlusion in 6.9% (12) and the rest of the etiologies were included in the category of other being only 1.1%(2). Visual acuity after Ahmed valve implantation had a median of 1.30 (0.70-2.3) in LogMAR. 59.2% (103) of the patients ended up using 3-4 hypotensive drugs after Ahmed valve implantation.

Those patients who presented hypertensive phase, had a higher HbA1c 8.3 (7.4-9.7) compared to patients without hypertensive phase 7.8 (6.5-9.1), this difference being statistically significant, p = 0.01 (figure 1).

**Figure 1.**
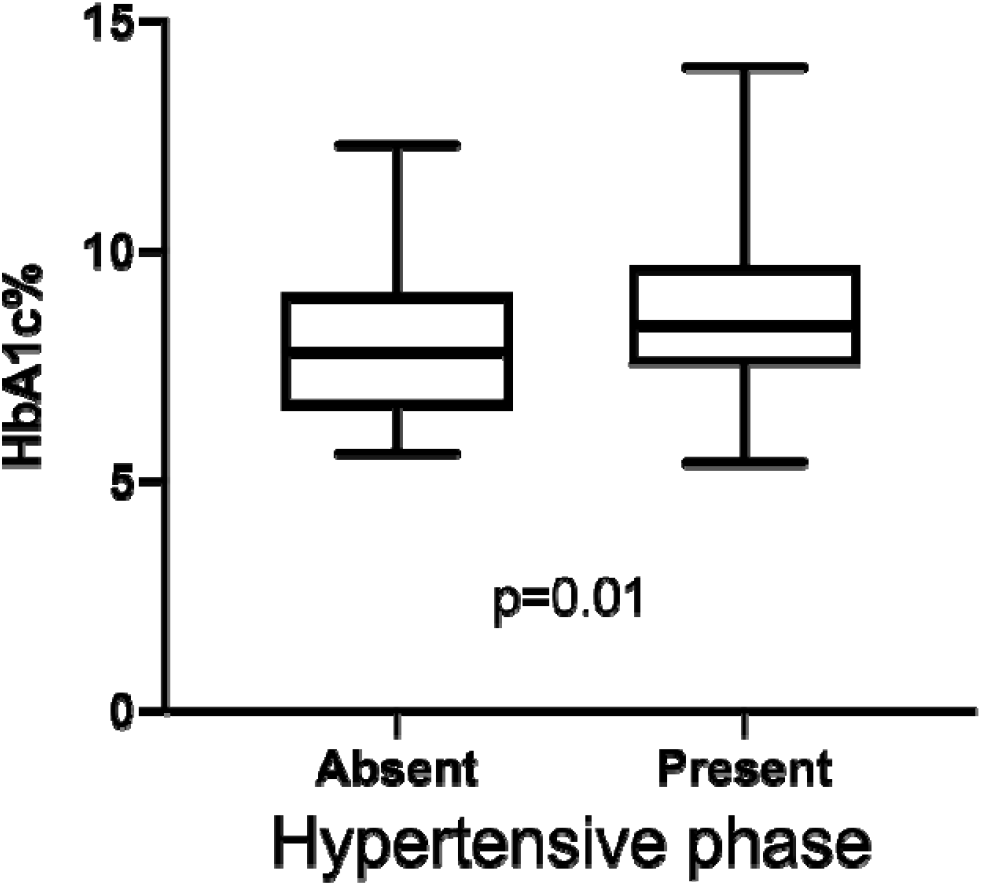
Box plot. Comparison of HB1Ac medians between groups. Mann–Whitney U test.

A difference (figure 2) was found in baseline IOP between patients who presented a hypertensive phase (47.84 ±11.6mmHg) compared to those who did not present a hypertensive phase (39.08 ±9.7mmHg), this difference was statistically significant, p<0.01.

**Figure 2.**
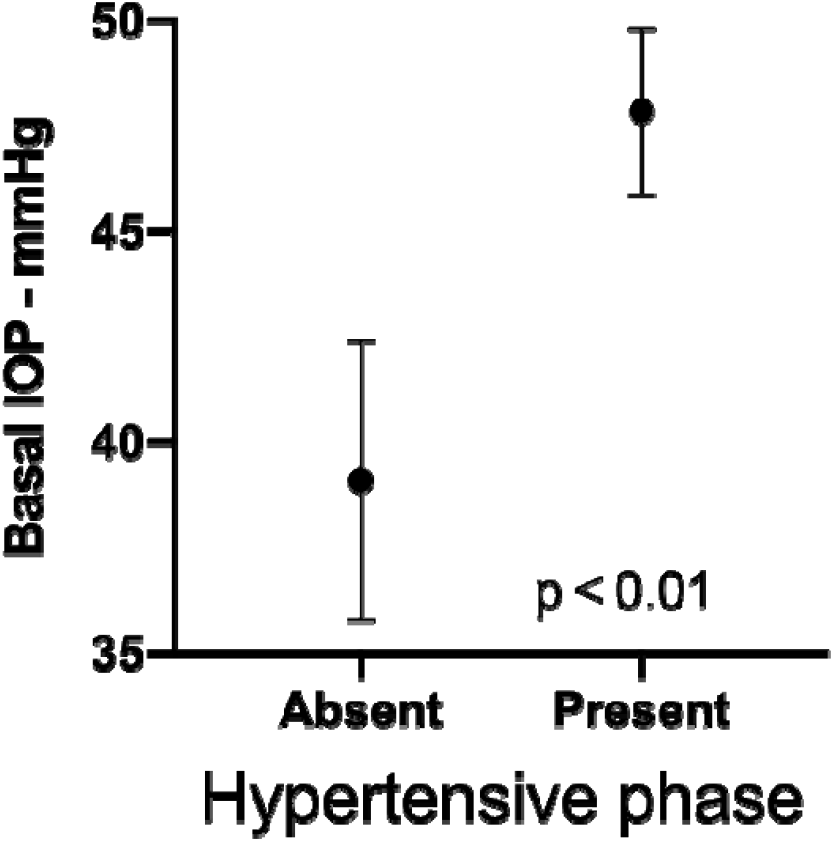
Error bar graph. Comparison of baseline intraocular pressure between groups. Student’s t-test.

The intraocular pressure with maximum topical treatment had a median of 35mmHg with IQR (26-42mmHg). This IOP measurement (figure 3) was higher in patients with hypertensive phase 35mmHg, IQR (28.0-43.0mmHg) than in patients without hypertensive phase 26mmHg, IQR (20-37.5mmHg), p<0.01.

**Figure 3.**
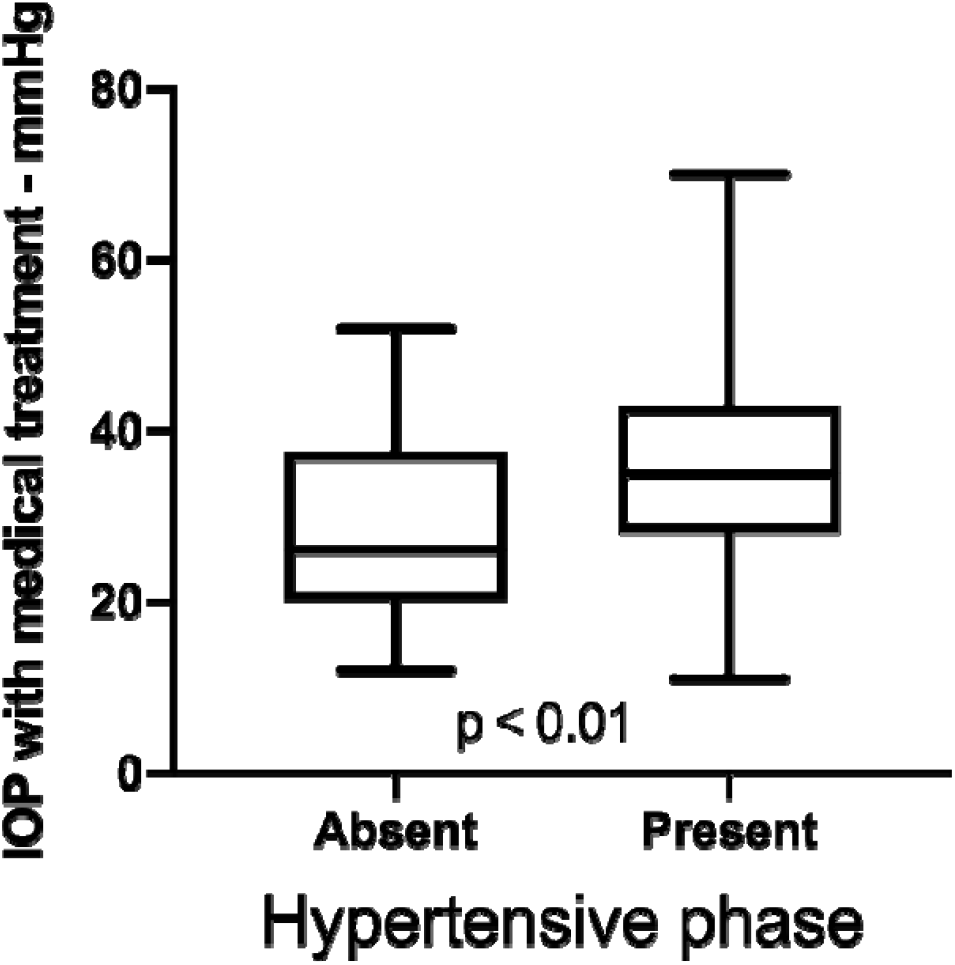
Box plot. Comparison of intraocular pressure with maximum topical treatment between groups. Mann–Whitney U test

Finally, Ahmed valve dysfunction had a statistically significant difference (p<0.01) in the frequency of Ahmed valve dysfunction among patients with hypertensive phase 65.9%(91) compared to patients without hypertensive phase 25.0%. (25). That is 2 out of 3 patients who presented post-surgical hypertensive phase presented dysfunction.

The Ahmed valve implant’s median survival (figure 4) was 8.9 months with a range of 3.7 to 14.2 months. The presence of a hypertensive phase (figure 5) marks a difference in survival between groups, patients with a hypertensive phase have a mean survival of 14.2 (11.5-16.9) months, while those patients who do not have a hypertensive phase is 37.8 (30.4-45.3) months. When analyzing the differences between the survival curves according to the maneuver, they show a significant difference with the Log-rank test (p<0.01).

**Figure 4.**
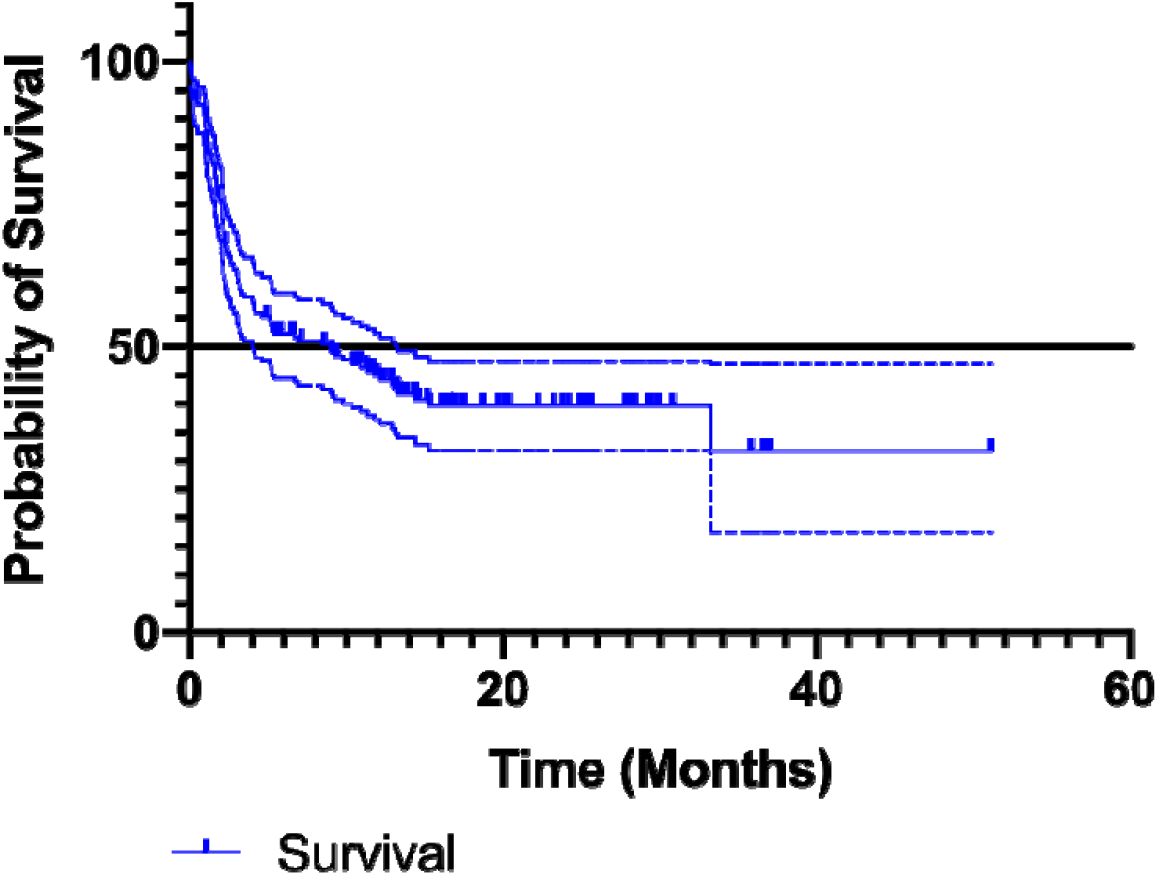
Overall survival curve of Ahmed valve implantation in patients with neovascular glaucoma. Median survival (solid line) and its 95% CI (dashed line) are shown. Median of survival 8.9 months.

**Figure 5.**
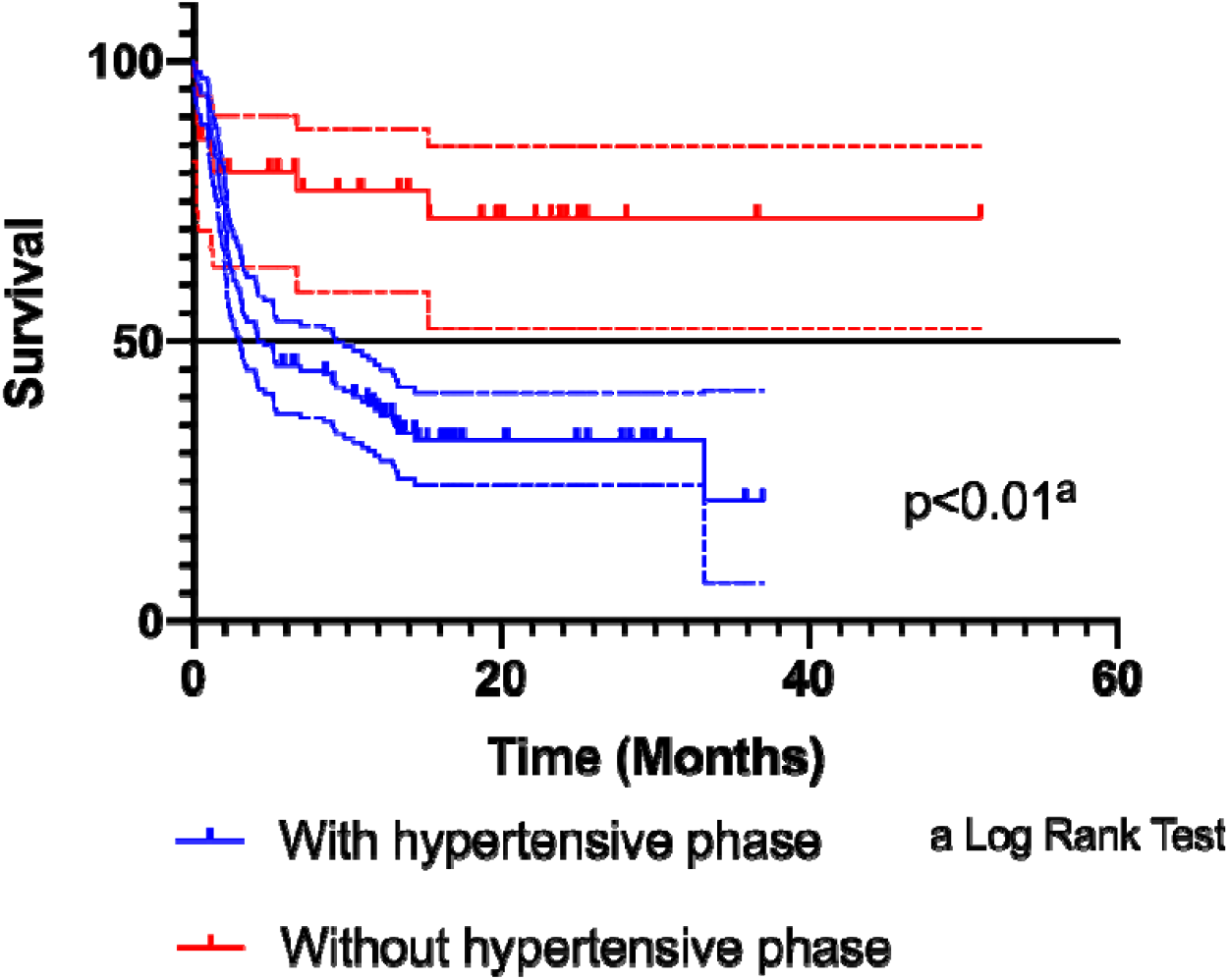
Survival curve of Ahmed valve implantation in patients with neovascular glaucoma comparing groups by the presence of hypertensive phase.

In the unadjusted Cox proportional hazards model (Table II), to determine risk factors associated with Ahmed valve failure, age under 50 years presents a hybrid risk (HR) of 1.54 (95% CI 1.04-2.30). The degree of neovascular glaucoma was not associated with an increased risk of failure, with an HR of 1.28 (95% CI 0.81-2.03). Glycosylated hemoglobin greater than 8% presents an increased risk for failure of 1.71 (95% CI 1.12-2.60).

**Table II.**
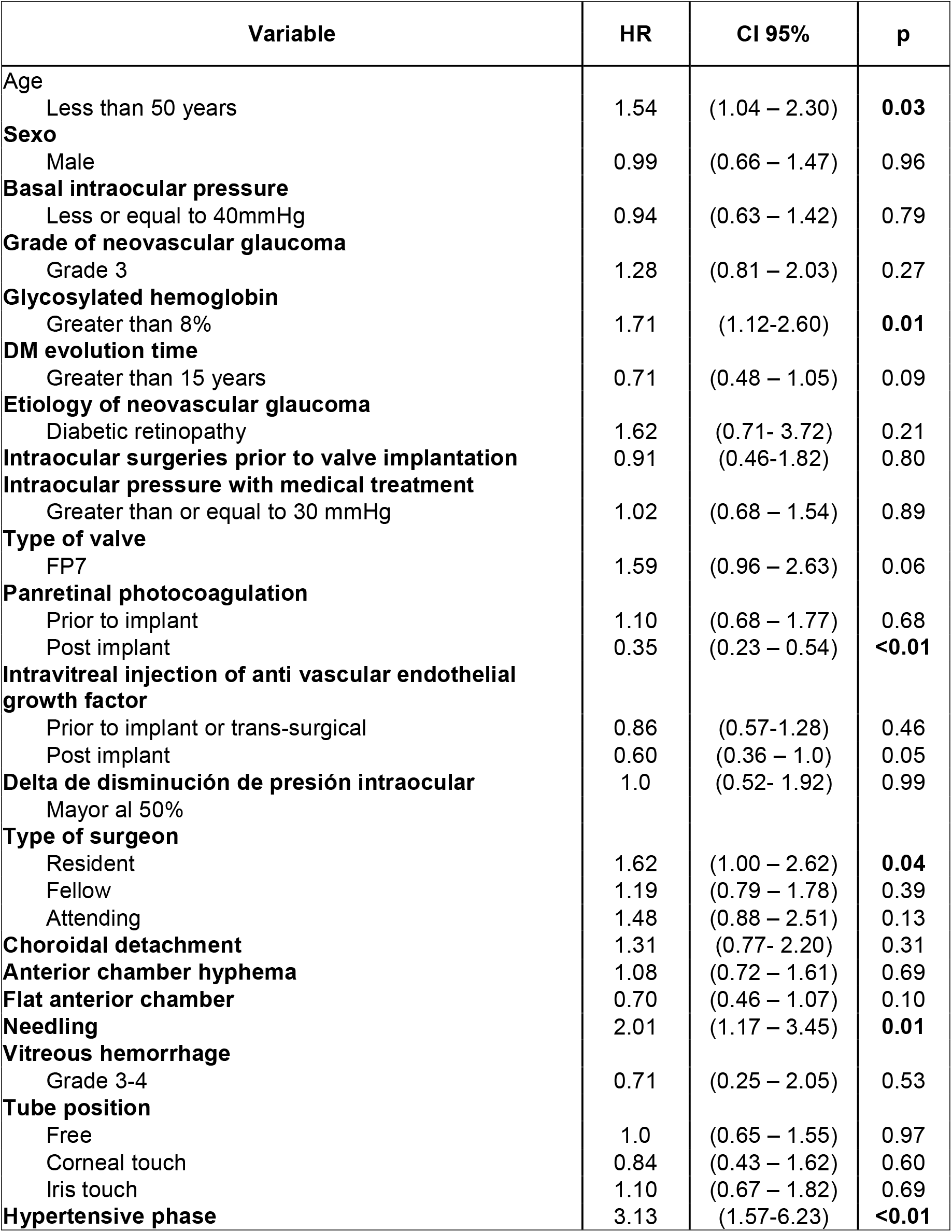
Cox proportional hazards model unadjusted for Ahmed valve failure. HR- Hazard Ratio; CI 95% - Confidence interval 95%

| Variable | HR | CI 95% | p |
| --- | --- | --- | --- |
| Age |  |  |  |
| Less than 50 years | 1.54 | (1.04 – 2.30) | **0.03** |
| **Sexo** |  |  |  |
| Male | 0.99 | (0.66 – 1.47) | 0.96 |
| **Basal intraocular pressure** |  |  |  |
| Less or equal to 40mmHg | 0.94 | (0.63 – 1.42) | 0.79 |
| **Grade of neovascular glaucoma** |  |  |  |
| Grade 3 | 1.28 | (0.81 – 2.03) | 0.27 |
| **Glycosylated hemoglobin** |  |  |  |
| Greater than 8% | 1.71 | (1.12-2.60) | **0.01** |
| **DM evolution time** |  |  |  |
| Greater than 15 years | 0.71 | (0.48 – 1.05) | 0.09 |
| **Etiology of neovascular glaucoma** |  |  |  |
| Diabetic retinopathy | 1.62 | (0.71- 3.72) | 0.21 |
| **Intraocular surgeries prior to valve implantation** | 0.91 | (0.46-1.82) | 0.80 |
| **Intraocular pressure with medical treatment** |  |  |  |
| Greater than or equal to 30 mmHg | 1.02 | (0.68 – 1.54) | 0.89 |
| **Type of valve** |  |  |  |
| FP7 | 1.59 | (0.96 – 2.63) | 0.06 |
| **Panretinal photocoagulation** |  |  |  |
| Prior to implant | 1.10 | (0.68 – 1.77) | 0.68 |
| Post implant | 0.35 | (0.23 – 0.54) | **<0.01** |
| **Intravitreal injection of anti vascular endothelial growth factor** |  |  |  |
| Prior to implant or trans-surgical | 0.86 | (0.57-1.28) | 0.46 |
| Post implant | 0.60 | (0.36 – 1.0) | 0.05 |
| **Delta de disminución de presión intraocular** | 1.0 | (0.52- 1.92) | 0.99 |
| Mayor al 50% |  |  |  |
| **Type of surgeon** |  |  |  |
| Resident | 1.62 | (1.00 – 2.62) | **0.04** |
| Fellow | 1.19 | (0.79 – 1.78) | 0.39 |
| Attending | 1.48 | (0.88 – 2.51) | 0.13 |
| **Choroidal detachment** | 1.31 | (0.77- 2.20) | 0.31 |
| **Anterior chamber hyphema** | 1.08 | (0.72 – 1.61) | 0.69 |
| **Flat anterior chamber** | 0.70 | (0.46 – 1.07) | 0.10 |
| **Needling** | 2.01 | (1.17 – 3.45) | **0.01** |
| **Vitreous hemorrhage** |  |  |  |
| Grade 3-4 | 0.71 | (0.25 – 2.05) | 0.53 |
| **Tube position** |  |  |  |
| Free | 1.0 | (0.65 – 1.55) | 0.97 |
| Corneal touch | 0.84 | (0.43 – 1.62) | 0.60 |
| Iris touch | 1.10 | (0.67 – 1.82) | 0.69 |
| **Hypertensive phase** | 3.13 | (1.57-6.23) | **<0.01** |

The presence of panretinal photocoagulation after implantation appeared as a protective factor for failure, with an HR of 0.35 (95% CI 0.23-0.54). The use of needling was also found to be a risk factor for failure with an HR of 2.01 (95% CI 1.17-3.45).

Finally, the presence of the hypertensive phase was the factor that presented a higher increased risk, with an HR of 3.13 (95% CI 1.57-6.23). The risk factors found in the bivariate model are represented graphically in Figure 6.

**Figure. 6.**
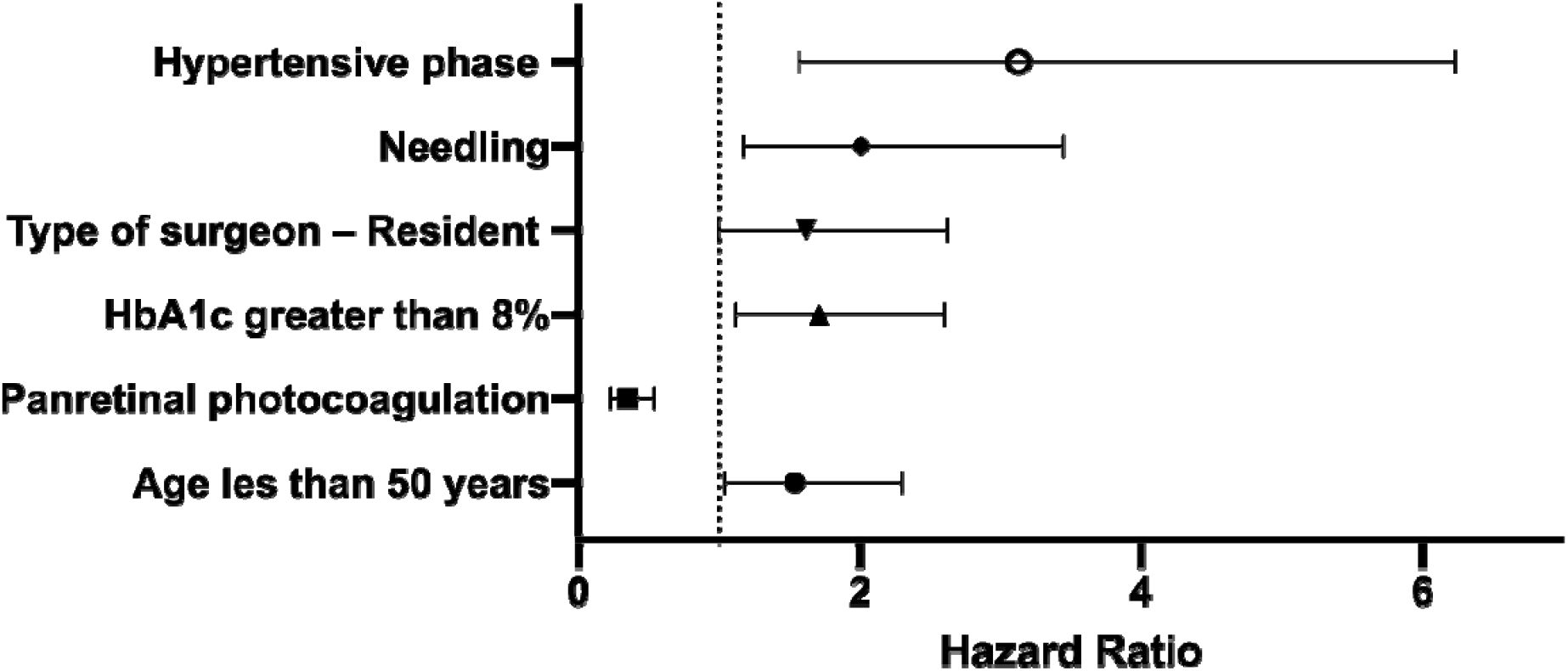
Forest plot of hazard ratio to present failure, Cox proportional hazards model unadjusted. HR with 95% CI are shown.

Regarding the adjusted Cox proportional hazards model (Table III), several models were carried out, starting with the variables that were significant in their unadjusted counterparts, finally, it was decided to create a parsimony model and leave those variables that best explained the outcome. Being that, the baseline intraocular pressure of less than 40mmHg presented a risk hybrid (HR) of 1.60 (95% CI 1.04-2.47), a percentage of glycosylated hemoglobin greater than 8% had an HR of 1.80 (95% CI 1.16-2.78), the type FP7 valve with an HR of 1.75 (95% CI 1.04 - 2.94), Resident as a type of surgeon had an HR of 1.67 (95% CI 1.02 - 2.73), the absence of panretinal photocoagulation after Ahmed valve implantation; HR 2.87 (95% CI 1.86-4.41) and the presence of hypertensive phase during follow-up had an HR of 3.24 (95% CI 1.60-6.59).

**Table III.** Multivariate Cox proportional hazards model for risk factors associated with Ahmed valve dysfunction.

| Variable | Multivariate model |  |  |  |
| --- | --- | --- | --- | --- |
|  | B | Hazard Ratio | CI 95% | p |
| Intraocular pressure | 0.47 | 1.60 | 1.04 – 2.47 | 0.03 |
| Glycosylated hemoglobin > 8% | 0.58 | 1.80 | 1.16 – 2.78 | <0.01 |
| Type of valve – FP7 | 0.56 | 1.75 | 1.04 – 2.94 | 0.03 |
| Type of surgeon – Resident | 0.51 | 1.67 | 1.02 – 2.73 | 0.03 |
| Panretinal photocoagulation post implant - Absent | 1.05 | 2.87 | 1.86 – 4.41 | <0.01 |
| Hypertensive phase - Present | 1.17 | 3.24 | 1.60 – 6.59 | <0.01 |

Model adjusted for: Baseline intraocular pressure, Glycosylated hemoglobin greater than 8%, Type of FP7 valve, Type of surgeon: Resident, Panretinal photocoagulation after valve implantation: Absent, Hypertensive phase: Present.

## Discussion

This study is the first, to our knowledge, that manages to present a multivariate model of the different factors that could influence the prognosis of these patients. The outcome of this surgery is blindness, so it is important to identify patients at higher risk.

In our study, we found that the proposed model identifies basal intraocular pressure greater than 40mmHg, glycosylated hemoglobin greater than 8%, type FP7 valve, surgery performed by a resident physician, the absence of photocoagulation after implantation, and the presence of a hypertensive phase. As the main risk factors associated with dysfunction. These findings are consistent with those reported in the literature, such as the work by Netland et al. which proposes age, the presence of Diabetes Mellitus, basal intraocular pressure, and the presence of neovascular glaucoma as risk factors associated with failure. However, only in the presence of neovascular glaucoma, the latter reached statistical significance (p <0.01) in the Cox proportional hazards model. Another limitation of this study was that it had a small sample, which probably prevents demonstrating the relationship with other variables. (16).

Similarly, Topouzis et al. present a second Cox proportional hazards model where the variables age, corneal transplant, basal intraocular pressure, race, the phakic status of the eye, and neovascular glaucoma as the risk factors associated with dysfunction. Like Netland, none of these factors were statistically significant. (17). Perhaps due to problems of lack of sample.

This work found a prevalence of hypertensive phase in patients with neovascular glaucoma who underwent Ahmed valve implantation of 79.3%, this figure is higher compared to that reported in the literature; in which reports range from 31% to 56% (18,19). On the other hand, the percentage of patients who did not resolve this hypertensive phase and later presented failure was found to be within the percentages reported in the literature, being 65.9% in our study compared to up to 76% reported by some authors (20). The presence of the hypertensive phase has been associated with many factors, including the type of valve, the type of glaucoma, and preoperative intraocular pressure, the latter being the most studied factor. Thus, a high basal intraocular pressure has been described as one of the predisposing factors for the development of the postoperative hypertensive phase (21). For all of the above, we can explain why our population presents a higher percentage of hypertensive phase, and it is that the basal intraocular pressure that we found was 46.03±11.8 mmHg, this value of intraocular pressure is higher by a wide margin than what is reported. in other studies (20–22).

Another clinical characteristic that we report in this study, which is not commonly reported, is the level of glycosylated hemoglobin, which we found in our patients, a median of 8.37% (7.18 - 9.70), which reflects poor systemic control. This is reflected in the causes of neovascular glaucoma since while in the literature the three most frequent causes (diabetic retinopathy, central retinal vein occlusion, and ischemic eye syndrome) are reported in percentages that vary between 33% (23)to 61.7%(24), our results show that in 92% of cases the most common cause in the Mexican population is diabetic retinopathy. With these data, we can infer that the Mexican population presents in more advanced stages of the disease, with worse metabolic and systemic control.

Ahmed valves can be mainly composed of two types of materials. Currently, the silicone model (FP7) is the most widely distributed worldwide and offers a more flexible valve body plate than the polypropylene model (S2). It is worth mentioning that both models have the same valve body area of 184mm^2^. Some studies have compared the results between both types of valves. In experimental models, the FP7 valve was associated with less inflammation than its S2 counterpart. (25). In this study, we found that the type of FP7 valve was a risk factor associated with Ahmed valve implant failure. These results contrast with what was reported by the workgroup of Ishida et al. (26), their results show that the FP7 valves have a greater IOP reduction, as well as a better success rate at three months and one year after implantation compared to the S2 model. This discrepancy in results may be due to the difference in sample size, since, in our study, model S2 is a small group. Additionally, our study included only patients with a diagnosis of NVG without previous valve implants and with a lower percentage of eyes that already had a previous surgical procedure.

To improve the survival of valve implants, the use of antimetabolites such as mitomycin C (MMC) and 5-fluorouracil (5-FU) has been tried. However, the results have not supported the routine use of these drugs. A study conducted by Al-Mobarak et al. found that the use of intraoperative MMC in S2 valves statistically significantly reduced valve implant survival at 2 years. This is believed to be because MMC induces fibrosis of the tissues around the valve body in pediatric patients. (27). On the other hand, Perez et al.(28) reported that the intraoperative use of MMC was associated with a lower prevalence of hypertensive phase 17.6% versus 55.0% in patients who did not use MMC, p=0.04.

Another alternative described in the literature to improve the prognosis of valve implants has been early management of the hypertensive phase with suppressors of aqueous humor production. Pakravan et al. randomized postoperative Ahmed valve implantation patients to early initiation of dorzolamide and timolol when intraocular pressure rose above 10mmHg versus conventional management. Their results showed that early initiation of hypotensive treatment was associated with a higher success rate, 63.2% versus 33.3%; p=0.008, as well as a lower percentage of hypertensive phase 23.4% vs. 66.0%; p<0.001 (29). Therefore, the use of hypotensive drugs early could be considered in those patients who undergo Ahmed valve implantation.

One of the strengths of this work is that it is the first study that has a multivariate model adjusted for the main confounding variables, as well as performing a stratified analysis of the results. Regarding the limitations of the study, we can mention that in the S2 valve subgroup we have few participants. The collection of data retrospectively implies the absence of observer bias, but it undoubtedly represents a limitation since we have to trust the data collected.

## Conclusion

Basal IOP less than 40mmHg, HbA1c >8%, type of FP7 valve, surgery performed by a resident, lack of photocoagulation after implantation, and the presence of a hypertensive phase are the independent risk factors for presenting dysfunction in the first postoperative year.

Early identification of the hypertensive phase can prolong the survival of the Ahmed valve.

## Data Availability

All data produced in the present work are contained in the manuscript

## Acknowledgement

This study is part of the masters studies at Sección de Estudios de Posgrado e Investigación. Escuela Superior de Medicina. Instituto Politécnico Nacional (IPN) of the master in science candidate Kingston Rodolfo Ureña-Wong.

